# Comparison of Radiologist-Assigned Categories and Quantitative Measures of Background Parenchymal Enhancement on Breast MRI

**DOI:** 10.1101/2020.03.09.20031153

**Authors:** Bethany L Niell, Mahmoud Abdalah, Olya Stringfield, Malesa Pereira, Dana Ataya, Yoga Balagurunathan, Robert Gillies, Natarajan Raghunand

## Abstract

Background parenchymal enhancement (BPE) on multi-parametric breast magnetic resonance imaging (mpMRI) reflects the volume and intensity of gadolinium uptake. We developed a semi-automated segmentation algorithm to extract and quantify measures of BPE and investigated the agreement of computed measures of BPE with radiologist-assigned categories. We retrospectively identified 123 high risk patients with breast mpMRI performed for screening indications. Pre- and post-gadolinium T1-weighted series with and without fat suppression were co-registered. Using Otsu’s method and an active contours method, the breast tissue was segmented from the chest wall and non-fat voxels were clustered to identify the amount of fibroglandular tissue. Median and inter-quartile ranges for the absolute volume of BPE (BPE cm3) and the percentage of BPE (BPE%) using a threshold PE of 30% were computed within the segmented FGT on the first post-contrast phase. Student’s t-test was used to evaluate BPE volume and BPE% by radiologist-assigned categories. Both BPE volume and BPE% differed significantly between minimal/mild and moderate/marked radiologist assigned categories (p=0.030 and 0.004, respectively). Using our newly developed semi-automated segmentation pipeline, we quantified fibroglandular tissue and BPE. The overlapping ranges of quantitative BPE corresponding to the radiologist-assigned BPE categories showcases the inter-reader variability with visual estimation of BPE.

## Introduction

Background parenchymal enhancement (BPE) on multi-parametric breast magnetic resonance imaging (mpMRI) reflects the volume and intensity of gadolinium uptake (1). Categorical measures of BPE are visually assessed by the breast radiologist and assigned to four categories: minimal, mild, moderate, and marked (1). Higher BPE categories increase the risk of breast cancer after adjustment for mammographic breast density or fibroglandular tissue (FGT) (2,3). Because categorical BPE assessments have a high degree of inter-reader variability, with reported kappas of 0.36-0.57 (3-6), development of quantitative BPE measures might be beneficial for incorporation into risk prediction models in the future.

Quantitative measures of enhancement could include any or all of the following: 1) average of the voxels of enhancement (PE = percent enhancement) above a pre-defined threshold, 2) total volume of tissue that enhances above the threshold value (absolute volume of BPE), and/or 3) the percentage of tissue enhancing above the threshold value relative to the total breast volume (BPE%). We developed a semi-automated segmentation algorithm to extract these quantitative measures of BPE and investigated the agreement of computed measures of BPE with radiologist-assigned categories.

## Materials and Methods

In this IRB approved HIPAA compliant study, we retrospectively identified 123 high risk patients with breast mpMRI performed for screening indications. The radiologist-assigned qualitative BPE category was extracted from the MRI report for each patient. A semi-automated segmentation pipeline was developed to calculate measures of FGT and BPE for each patient. One pre-gadolinium T1-weighted non-fat-suppressed (T1w-NFS), one pre-gadolinium and four post-gadolinium T1-weighted fat-suppressed sequences were co-registered to create the initial mask which included thorax, chest wall, pectoralis muscle, breast, and surrounding fat. Using the initial mask, the caudal and cephalad extent of the breasts, as well as the mid-sternal intersection of both breasts, were demarcated in the axial and coronal planes, respectively. A refined chest mask was obtained by Otsu’s method from the Pre-T1w-FS sequence (7). Chest components were merged using an active contours method (8). Final total breast volume (TBV) was generated by removing the chest mask from the initial mask. The pre-contrast T1w-NFS sequence was used to exclude fat. The remaining (non-fat) voxels were clustered using Otsu’s method to identify voxels containing FGT (Figure 1).

**Figure 1.**
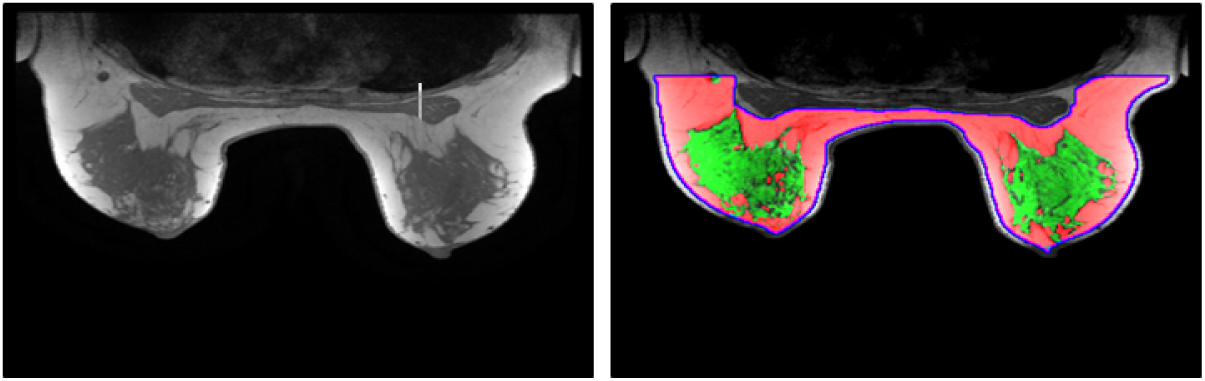
Segmented breast tissue volume (pink) and FGT volume (green)

Median and inter-quartile ranges for absolute volume of BPE and BPE% using a threshold PE of 30% were computed within the segmented FGT on the first post-contrast phase. Student’s t-test was used to evaluate BPE volume and BPE% by radiologist-assigned categories.

## Results

Of the 123 cases, 46 (37%) were assigned a minimal BPE category, 41 (33%) a mild BPE category, 25 (20%) a moderate BPE category, and 11 (9%) a marked BPE category by the radiologist. Median and inter-quartile ranges for the volume of BPE (measured in cubic centimeters) by radiologist-assigned category are shown in Table 1.

**Table 1.** Median and interquartile ranges for BPE volume (cm^3^) by BPE category

| **Radiologist-assigned**<br>**BPE category** | **BPE volume in cm<sup>3</sup>**<br>**Median (Interquartile range)** |
| --- | --- |
| Minimal | 57.2 (24.4-100.1) |
| Mild | 41.8 (30.7-65.4) |
| Moderate | 70.6 (43.5-111.1) |
| Marked | 67.1 (54.3-137.9) |

The percentage of tissue enhancing above the 30% threshold value relative to the total breast volume (BPE %) was also quantitatively extracted. BPE% median and inter-quartile ranges by radiologist-assigned category are shown in Table 2.

**Table 2.**
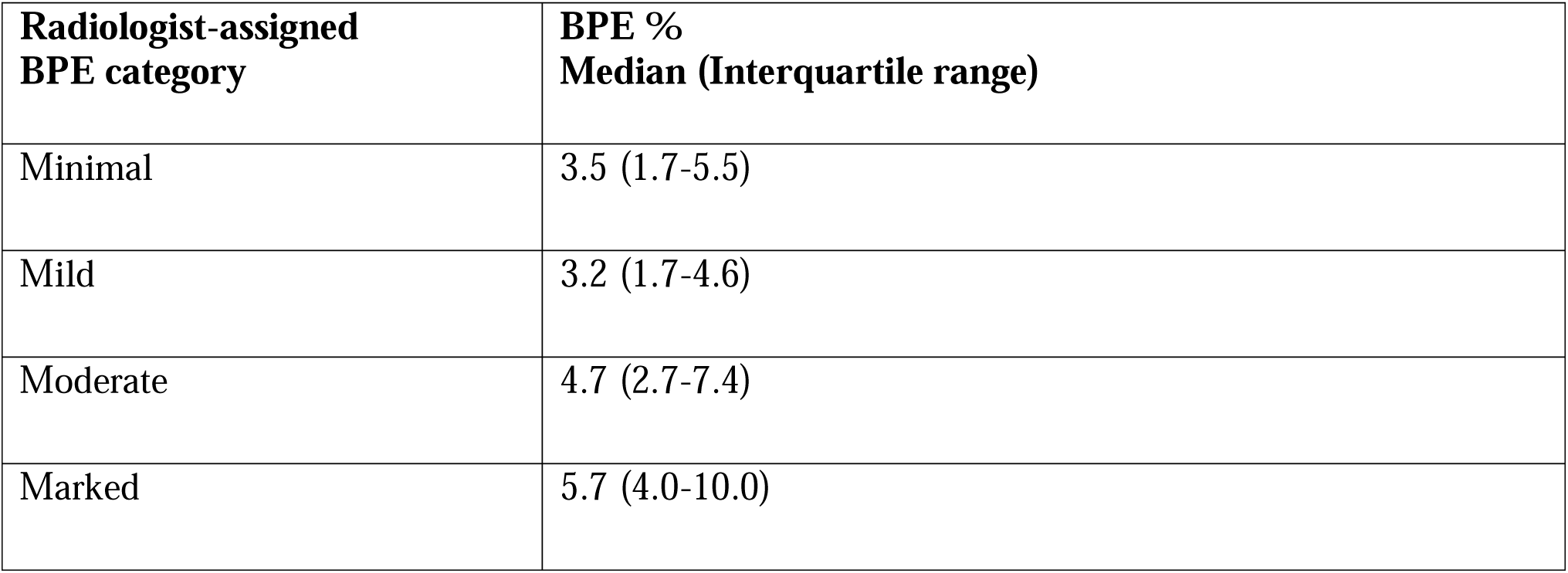
Median and interquartile ranges for BPE% by BPE category

Both BPE volume and BPE% differed significantly between minimal/mild and moderate/marked radiologist assigned categories (p=0.030 and 0.004, respectively). Figure 2 displays BPE% by radiologist assigned categories.

**Figure 2:**
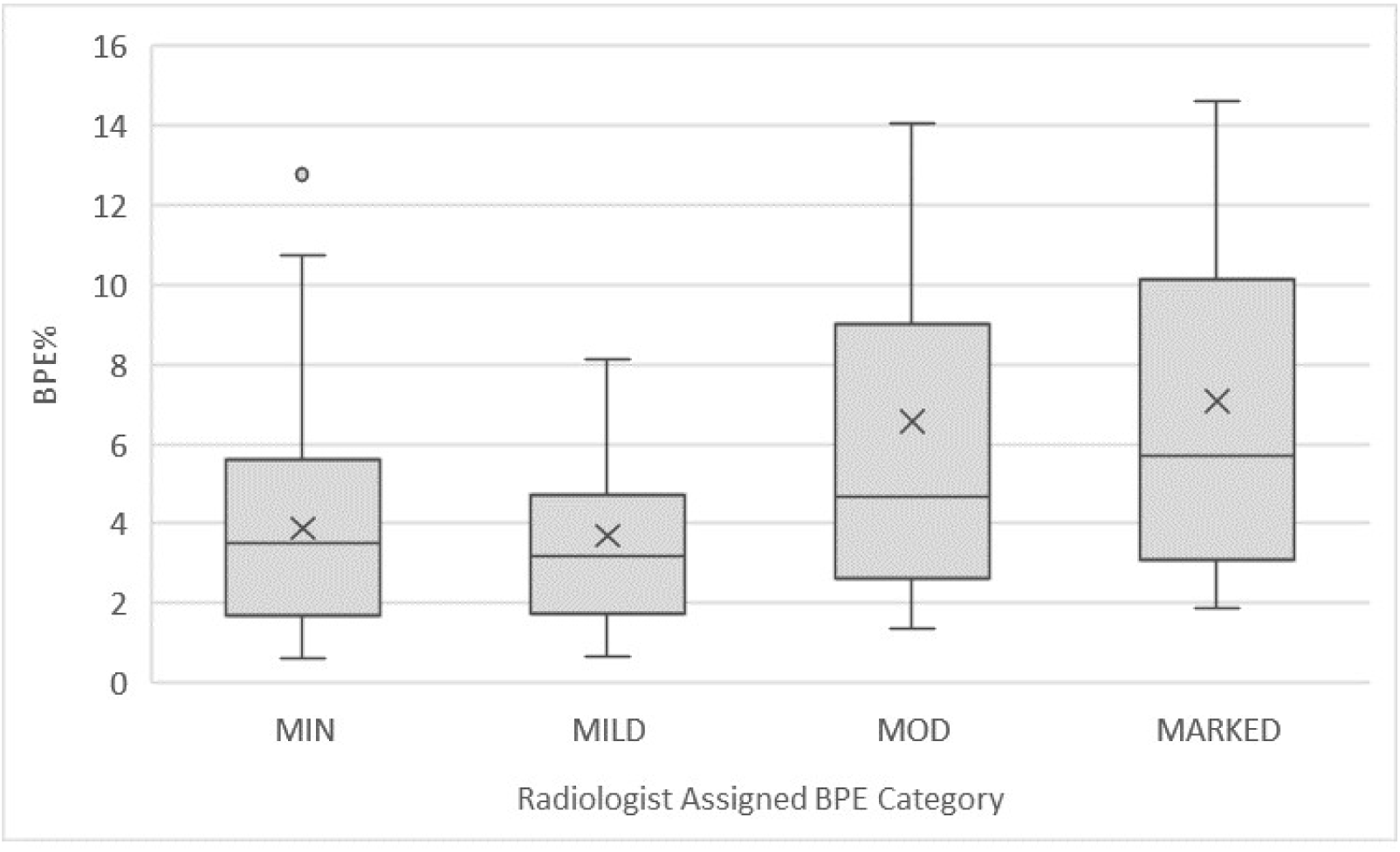
Box plot of BPE% by BPE category

## Discussion

Our novel semi-automated segmentation pipeline was developed to quantify TBV, FGT, and BPE. In our cohort, quantitative measures of BPE% and BPE volume (cm^3^) differed significantly between moderate/marked and minimal/mild radiologist-assigned categories. The overlapping ranges of quantitative BPE corresponding to the radiologist-assigned BPE categories showcases the inter-reader variability with visual estimation of BPE. T Because background parenchymal enhancement at breast MRI is an imaging biomarker for breast cancer risk, this inter-reader variability in BPE categorical assessments highlights the need for development and validation of quantitative measures as an important intermediate step towards incorporation of BPE into future risk prediction models.

Our study has limitations. Segmenting the breasts from the chest wall was a semi-automated process and may be subject to operator dependency. In addition, this was a single-institution study, so determination of quantitative BPE measures was based on a single breast MRI protocol. Additional studies utilizing a larger cohort with varied institutional protocols would be useful in establishing generalizability.

In summary, BPE is now recognized as an important imaging biomarker of breast cancer risk. Our novel semi-automated segmentation pipeline permits quantification of BPE. In the near future, we hope that quantification of BPE will allow for standardization of BPE reporting and assessment, so those BPE measures can be incorporated into breast cancer risk prediction models.

## Data Availability

Data would be available upon request following publication.

